# Acceptability of a proposed new tuberculosis vaccine among people deprived of liberty in Brazil

**DOI:** 10.64898/2025.12.02.25341289

**Authors:** José Victor Bortolotto Bampi, Ghislaine Gonçalez de Araujo Arcanjo, Karina Marques Santos, Michele Souza Ventura, Rebecca A. Clark, Katherine A. Thomas, Richard G. White, Julio Croda

## Abstract

**Background:** Globally tuberculosis (TB) persists as a public health threat, with an unequal burden of disease among populations. Targeting disease control, new vaccine candidates for adults and adolescents are currently in the development pipeline. If available, these products can be directed in vaccination campaigns for vulnerable populations, such as people deprived of liberty (PDL). This study aims to assess acceptability towards a proposed new TB vaccine and factors associated with hesitancy in a carceral settings at eight prison units in Brazil.

**Methods:** We performed a cross-sectional study among PDL in six male and two female prison units within 6 Brazilian cities from April 2025 to October 2025. Eligible participants included adults (over 18 years old) who could provide consent. Through prison census, we randomly selected 130 individuals for initial evaluation with structured questionnaire about sociodemographic status, TB disease beliefs, willingness to receive a hypothetical new TB vaccine, sources of information trusted and select reasons that could make them not take a vaccine. We also requested that individuals rated affirmations regarding TB vaccines in the Likert scale. We compared participants characteristics and answers to vaccine questionnaire within prison gender and reported vaccine acceptance groups.

**Results:** Of the 945 individuals evaluated, 4 were excluded due to missing questionnaire results, with 941 included for main analysis. In total, 95.2% of individuals reported that they would take the TB vaccine if available for them, with 94.1% acceptance in male prisons and 98.7% in female prisons. Compared to females, male individuals reported more distrust in vaccine safety (28.9% vs 14.5%, p <0.001), more community coercion to vaccine uptake (15.4% vs 5.6%, p<0.001) and worse TB knowledge (44.6% vs 31.2%, p<0.001). Overall, among individuals that would not accept vaccination, 77.8% and 55.6% of them did not trust vaccine safety and efficacy, respectively, 60.0% did not trust healthcare workers and 20.0% reported community coercion.

**Conclusion:** We found that acceptability of a new TB vaccine in Brazilian prisons was very high. Despite few differences in intent to vaccinate regarding gender, individuals that refused vaccination more often reported problems with vaccine and healthcare trust, as well as a significant proportion of them reported possible community coercion for vaccine uptake. Our findings suggest that a new TB vaccine would be well accepted among PDL.

## Background

Globally tuberculosis (TB) persists as the leading cause of death from a single infectious agent, affecting over 10 million people in 2025 (1). Within the End TB Strategy goals there are many proposed approaches to face this global health threat (2), amongst them, new vaccines that can provide protection against *Mycobacterium tuberculosis* and the development of TB disease in adolescents and adults would be major scientific breakthroughs for disease control across all populational groups(3).

Despite that, there is no current vaccine product licensed for TB protection in adults and adolescents but there are over a dozen candidates in the development pipeline using different platforms (4), many of those already in phase 2 and 3 clinical studies as the most recent recombinant protein vaccine M72/AS01_E–4_ the live-attenuated MTBVAC candidates (5,6). Within this thriving reality of possibilities for interventions trials and future immunization campaigns, it is paramount that we explore vaccine acceptability, hesitancy and overall perception of these circumstances amid the targeted populations.

Currently, TB incidence rates in prison settings are among the highest recorded around the globe (7) and can act as reservoirs of disease burden, driving the current incidences in the overall community (8,9). In Latin America, previous studies have shown that the increasing incarceration rate in the last decade is one of the main factors for the increased TB incidence overall, amounting to TB rates 26 times higher in PDL compared with the national averages (10,11). Targeted vaccination campaigns for PDL could add substantially to TB control nationally, thus, it is essential that such a vulnerable population is assessed for willingness to participate in interventions that could reduce the burden of disease.

Most available studies evaluating vaccine acceptancy in prisons are related to high income countries or COVID-19 vaccines (12–14). The reported coverage and acceptance are variable; however, barriers for coverage as distrust in vaccine products safety and efficacy, distrust in prison staff members and healthcare workers are commonly reported in those studies. In Brazil, despite recent decreases in coverage rates of Measles, Diphtheria-tetanus-pertussis and Poliomyelitis vaccines, vaccination has historical high acceptance in the overall population (15,16). Yet evidence regarding PDL is scarce; reports from government agencies (17) and individuals states (18) indicate that acceptability of COVID-19 vaccines was high within prisons, however there is no official country level data.

Nevertheless, factors leading to vaccine uptake are complex and have variable determinants amidst populations, vaccine products and targeted diseases (16,19); meaning that current data might not reflect acceptability for a new TB product. Elements such as TB stigma within prisons, individual risk perception, trust in healthcare workers and information sources (20–22) might unpredictably influence immunization mobilizations success.

On account of these knowledge gaps, this study aims to evaluate acceptability and hesitation to a new hypothetical TB vaccine, as well as TB beliefs and knowledge, through a survey performed in 6 male and 2 female prisons across 6 cities in Brazil.

## Methods

### Study setting and design

We conducted a cross-sectional study at six male prison units located within six cities (Belo Horizonte, Campo Grande, Manaus, Porto Velho, Salvador and Santa Cruz do Sul) and two female prison units located within two cities (Campo Grande and Manaus) in Brazil, covering all country regions. All units are adult (over 18 years old) closed system prisons, where individuals do not leave prison during incarceration. Data collection and study procedures occurred between April 2025 and October 2025.

### Data collection

Participant selection was performed first with a random selection of 130 PDL. We obtained a complete prison unit census from each unit in the week prior to study procedures and performed a complete randomized selection of potential participants based on name and prison identifiers. The initial sampled individuals were approached by study personnel and inquired about willingness to participate in study procedures. Due to carceral transferences, prison security setting and time constraints, study personnel could not approach the maximum number of individuals included in all prisons. All of those who were willing and provided informed consent were interviewed by a trained study researcher that collected answers to questions in a structured sociodemographic and vaccine acceptability questionnaire in paper, that was later transferred to electronic format. After data collection and sample processing, we excluded individuals with incomplete questionnaire answers recorded from the main analysis (**Figure 1**).

**Figure 1.**
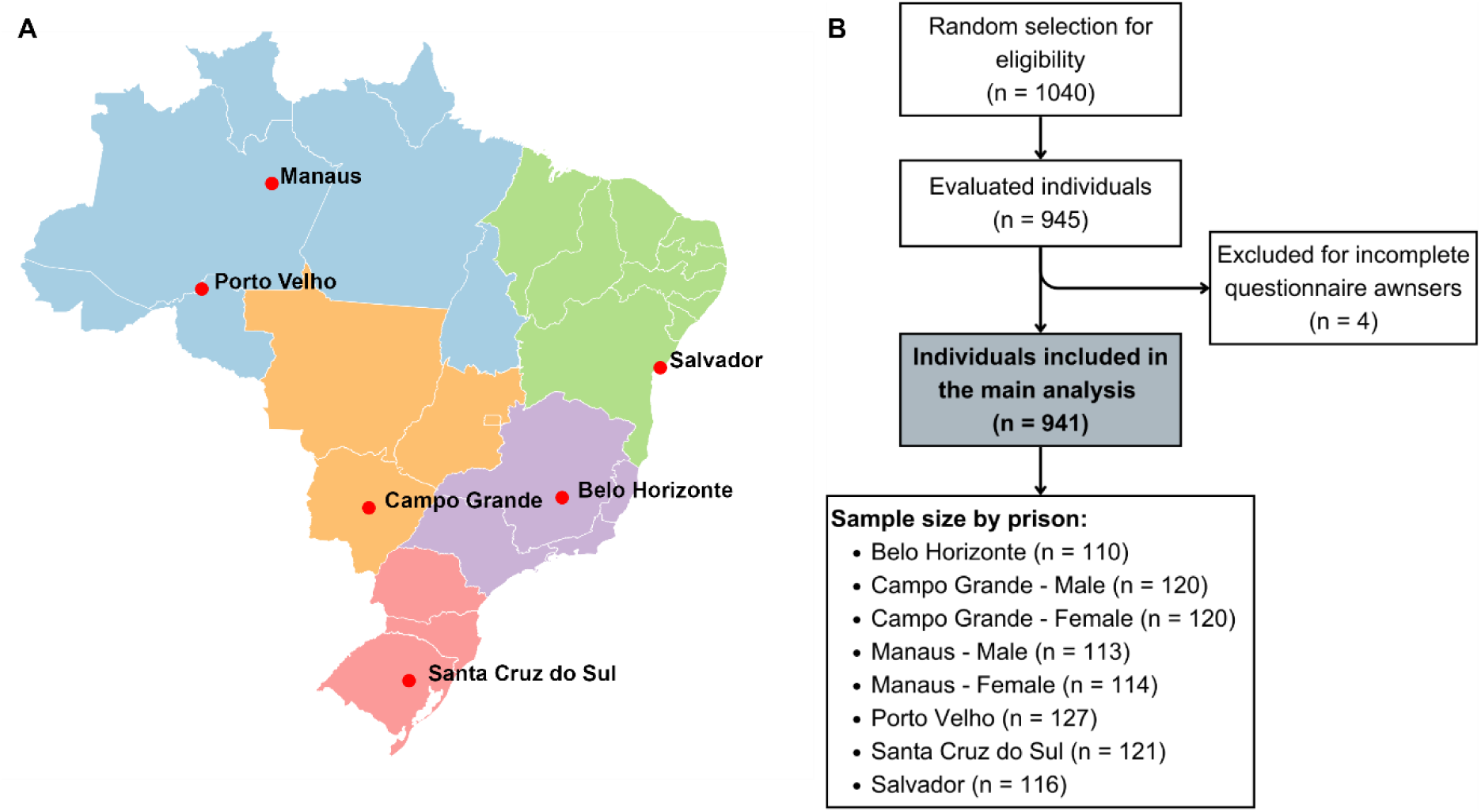
Cities included in the study evaluation and flowchart of inclusion of study participants. **A –** Cities where the study prison units were located within Brazil. Five different country regions are represented in different colors, at least one prison was evaluated in each region. **B –** Flowchart of participant inclusion and number of participants at each site.

### Vaccine acceptability questionnaire

Participants were inquired about vaccine acceptability through a standardized questionnaire (**Supplementary materials, Appendix 1**). Initially, study investigators questioned directly about prior TB knowledge and, utilizing the Likert scale (“Strongly agree”, “Agree”, “Don’t know”, “Disagree” and “Strongly disagree”), requested participants to assess affirmations about TB disease concerns.

Currently, since there are no licensed TB vaccines in use for the adult population, study investigators solicited that participants would think about a scenario were a new vaccine was available for them, affirming: “If a new vaccine for adolescents/adults was approved for tuberculosis (TB) and recommended by the Brazil Ministry of Health, how strongly do you agree or disagree with the following affirmations?”. This statement was followed up by affirmations assessing vaccine trust and healthcare trust. After that, individuals were questioned directly about the decision to take the possible vaccine, “If a new vaccine for adolescents/adults was approved for tuberculosis (TB), available to you today, recommended by the Brazil Ministry of Health, and your healthcare provider recommended it, would you choose to get the vaccine?”. Interviewed individuals could choose between the answers “Yes”, “No” and “Unsure”; for analysis purposes we considered acceptance as those who answered “Yes” and aggregated the “No / Unsure” answers as hesitant. Finally, individuals were asked to select options from a multiple-choice list of what they considered trustworthy sources of information for TB vaccine recommendations and other reasons they might consider not taking a TB vaccine if it were available.

### Indicators of possible vaccine hesitancy

We decided to utilize the 5C vaccine hesitancy model (23) to approach possible drivers of vaccine denial in future realistic vaccination campaigns scenarios. Through a composite analysis of answers provided, we assessed six categories of possible obstacles to vaccine uptake within the 5C model.

**Confidence** was evaluated through distrust in vaccine (efficacy and safety) and healthcare system. Composite distrust in vaccine safety was considered present if individuals answered “Disagree” or “Strongly disagree” to “I think a new tuberculosis (TB) vaccine would be safe” or if they reported “Health-related barriers”, “Pregnancy or lactating barriers”, “Distrust in vaccines in general” or “Distrust in TB vaccines” among reasons to not take the vaccine. Composite distrust in vaccine efficacy was considered present if individuals answered “Disagree” or “Strongly disagree” to “I think a new tuberculosis (TB) vaccine would be effective” or if they reported “Distrust in TB vaccines” among reasons to not take the vaccine. Composite distrust in healthcare providers and workers was considered present if individuals answered “Disagree” or “Strongly disagree” to “ I would trust the health workers who would give me a new tuberculosis (TB) vaccine”, if they not selected “Facility-based healthcare workers or community/lay healthcare worker (health care professionals)” or “Ministry of Health or other government officials (government agencies)” as trustworthy sources of information or if they selected “I do not trust the healthcare system and/or the healthcare workers” in respect of reasons to not take the vaccine.

**Complacency** was assessed as concerns about TB disease, we considered it present if individuals answered “Agree” OR “Strongly Agree” to the affirmations “I am concerned that I could develop TB” or “If I developed TB, I am concerned that I would become seriously ill or die” and did not select the option “TB beliefs (e.g., I am not concerned about developing TB)” in the reasons they might not take the vaccine.

**Calculation** was evaluated as insufficient TB knowledge if individuals did not report previous TB treatment or if they answered “I had heard of TB and did not knew much about it” or “I have never heard of TB before” to the question “Before today, how much did you know about tuberculosis (TB)?”. We also considered as not adequate information sources if individuals did not select “Facility-based healthcare workers or community/lay healthcare worker (health care professionals)” or “Ministry of Health or other government officials (government agencies)” as trustworthy sources of information.

Although we did not questioned individuals directly about Constrains of access to the vaccine or Collective responsibility about community transmission, we considered a composite variable of Collective constrains, where these individuals indicated that community factors would influence their informed decisions to take or not the vaccine. This was considered present if participants selected “Family or community-related barriers”, “Fear of repression from other people deprived of liberty” or “I don’t believe that i would have the option to deny a TB vaccine, even if i did not wanted to take it” in the reasons they might not take the vaccine.

### Data management and statistical analysis

All data collected was stored in an online RedCap® (Vanderbilt University, Tennessee, USA) database hosted by Fiocruz, Mato Grosso do Sul, Brazil. Data analysis was performed utilizing R statistical software (24). We utilized Chi-square and Fisher tests for categorical variables to determine differences in the response variables among subgroups.

## Results

During the study period, 945 individuals were evaluated through study procedures from 1040 PDL selected previously. Initially, we excluded 4 individuals with incomplete questionnaires, amounting to a final sample of 941 individuals from the 8 prisons included in the main analysis **(Figure 1).** Overall, 75.1% of PDL were from male prisons, the median age was 32 years old (IQR 27-39) and most were mixed-race (58.0%). More frequently, individuals were single (59.4%) had incomplete basic education (48.4%) and had a total family income up to 2 minimum wages (62.2%). Current incarceration time was less than a year for around one third (37.9%) of participants and most (71.9%) had a history of previous incarceration. We encountered a high prevalence of self-reported alcohol use disorder (4.5%), illicit drug use (15.3%) and current tobacco use (25.8%), however reported HIV positive status was low (1.4%), similar to the overall Brazilian population estimates (25). The proportion of previous TB treatment reported was considerable at 9.7% and 34.3% reported previous contact with a positive TB individual in the same cell. Male and female prisons had substantial differences in the previous reported characteristics, but the proportion of HIV positive status, previous TB history and family income did not statistically differ between the groups **(Table 1)**.

**Table 1.**
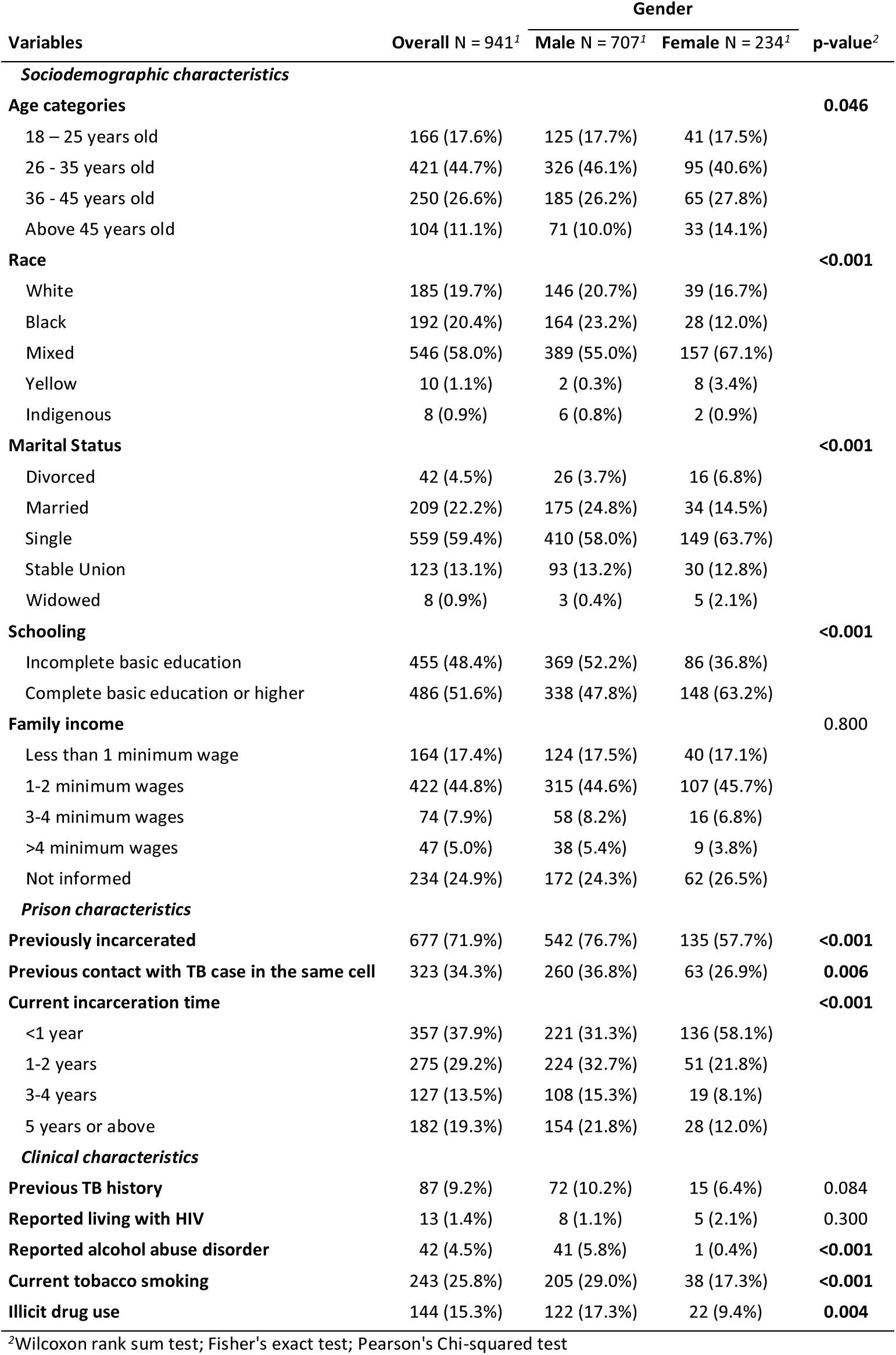
Characteristics of incarcerated individuals included in the vaccine acceptability study, by gender (n = 941)

Most of the participants interviewed (95.2%) in all prisons reported that they would take the new vaccine if available to them, only three (6.6%, 3/45) of participants that said they would not accept it were from female prisons. Participants that would decline a new TB vaccine were very similar to those who would accept a vaccine regarding sociodemographic and prison characteristics. However, we identified that those that would decline a vaccine more frequently reported tobacco smoking (44.4% vs 24.9% p=0.003) and current drug use (26.7% vs. 14.7%, p = 0.030). (**Supplementary Table 1**).

Participants that would accept a vaccination had very low disagreement rates with answers related to confidence in the Likert scale answers. (**Figure 2**). We also identified that trust in healthcare workers (64.4% vs 81.6%, p=0.004), the Brazilian government (42.2% vs 58.8%, p = 0.028) and community leaders (0% vs 8.0%, p=0.042) was lower in those who refused the vaccine. Those individuals refusing the hypothetical vaccine also reported more causes related to health barriers (31.1% vs. 13.5%, p<0.001) to not accept it, distrust was also significantly more frequently reported by the same group, more importantly regarding distrust in general vaccines (28.9% vs. 5.8% p<0.001), TB vaccines (37.8% vs 5.0%, p < 0.001) and healthcare workers (8.9% vs 1.2%, p = 0.004) **(Table 2).**

**Figure 2.**
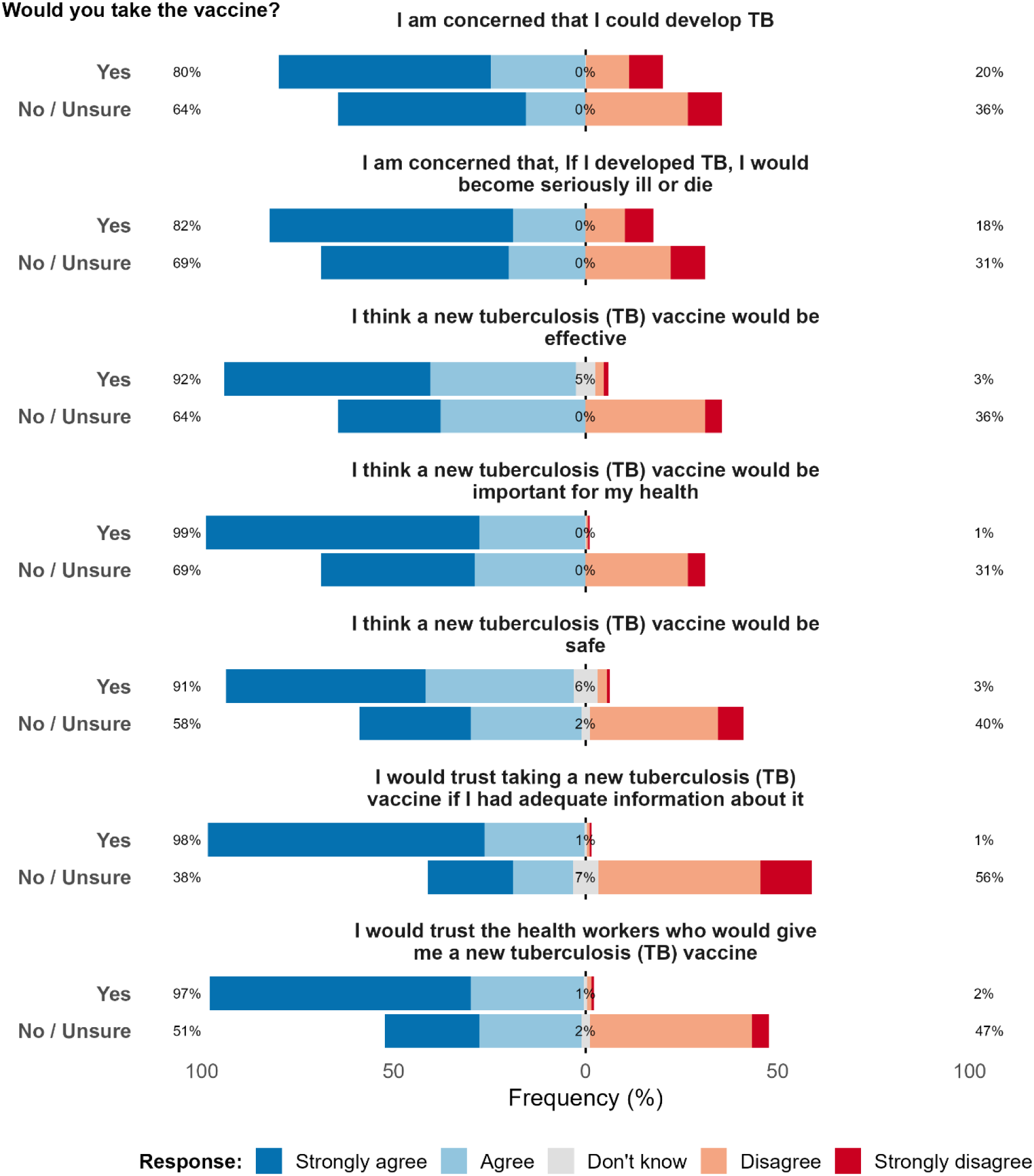
Likert scale responses to questions related to tuberculosis concerns and vaccine acceptability among people deprived of liberty, stratified by willingness to take a proposed TB vaccine (N = 941)

**Table 2.** Vaccine acceptability questions assessing knowledge, information sources and reasons to not get vaccinated in people deprived of liberty, by willingness to take a proposed TB vaccine (n = 941)

| Variables | Overall<br>N = 941<sup>1</sup> | Would you take the TB vaccine? |  | p-value<sup>2</sup> |
| --- | --- | --- | --- | --- |
|  |  | No / Unsure<br>N = 45<sup>1</sup> | Yes<br>N = 896<sup>1</sup> |  |
| **"Before today, how much did you know about tuberculosis (TB)?"** |  |  |  | **0.800** |
| I had heard of TB, and knew a lot about it | 189 (20.1%) | 8 (17.8%) | 181 (20.2%) |  |
| I had heard of TB, and knew some things about it | 364 (38.7%) | 19 (42.2%) | 345 (38.5%) |  |
| I had heard of TB, but did not know much about it | 342 (36.3%) | 15 (33.3%) | 327 (36.5%) |  |
| I had never heard of TB | 46 (4.9%) | 3 (6.7%) | 43 (4.8%) |  |
| **"Has someone you know ever had tuberculosis (TB)?"** |  |  |  | **0.049** |
| Yes, someone I know has had or currently has TB | 594 (63.1%) | 30 (66.7%) | 564 (62.9%) |  |
| No, I do not know anyone who has had TB | 329 (35.0%) | 12 (26.7%) | 317 (35.4%) |  |
| Unsure | 18 (1.9%) | 3 (6.7%) | 15 (1.7%) |  |
| **What sources of information would you trust the most to recommend a new TB vaccine?** |  |  |  |  |
| Facility-based healthcare workers or community/lay healthcare worker (health care professionals) | 760 (80.8%) | 29 (64.4%) | 731 (81.6%) | **0.004** |
| Family, friends, colleagues (personal network) | 369 (39.2%) | 13 (28.9%) | 356 (39.7%) | 0.150 |
| Ministry of Health or other government officials (government agencies) | 546 (58.0%) | 19 (42.2%) | 527 (58.8%) | **0.028** |
| Religious leaders (faith) | 140 (14.9%) | 3 (6.7%) | 137 (15.3%) | 0.110 |
| Other community leaders (community-based network) | 72 (7.7%) | 0 (0%) | 72 (8.0%) | **0.042** |
| Social media/internet sources | 77 (8.2%) | 1 (2.2%) | 76 (8.5%) | 0.200 |
| Other | 12 (1.3%) | 4 (8.9%) | 8 (0.9%) | **0.002** |
| **What are some reasons you might not get a TB vaccine if it were available to you?** |  |  |  |  |
| Health-related barriers (e.g. allergies or other health conditions) | 135 (14.3%) | 14 (31.1%) | 121 (13.5%) | **0.001** |
| Pregnancy or lactating barriers (e.g., safety while pregnant or breastfeeding) | 12 (1.2%) | 1 (3.0%) | 11 (1.2%) | 0.150 |
| Family or community-related barriers (e.g., fear of reaction/ stigma) | 64 (6.8%) | 4 (8.9%) | 60 (6.7%) | 0.500 |
| TB beliefs (e.g., I am not concerned about developing TB) | 9 (1.0%) | 2 (4.4%) | 7 (0.8%) | 0.065 |
| Distrust in vaccines in general (e.g., negative experiences with vaccines, I do not like injections, I am concerned about potential side effects of vaccines [e.g., long-term or short-term side effects including fertility]) | 65 (6.9%) | 13 (28.9%) | 52 (5.8%) | **<0.001** |
| Distrust in TB vaccines (e.g., TB vaccine is not important or necessary, I do not think a new TB vaccine will work or stop me from getting TB, I do not think a new TB vaccine will be safe, or I am concerned about potential side effects of a new TB vaccine) | 62 (6.6%) | 17 (37.8%) | 45 (5.0%) | **<0.001** |
| I do not trust the healthcare system and/or the healthcare workers | 15 (1.6%) | 4 (8.9%) | 11 (1.2%) | **0.004** |
| Fear of repression from other people deprived of liberty | 14 (1.5%) | 1 (2.2%) | 13 (1.5%) | 0.500 |
| I don't believe that i would have the option to deny a TB vaccine, even if I did not wanted to take it | 59 (6.3%) | 5 (11.1%) | 54 (6.0%) | 0.200 |
| None of the above | 625 (66.4%) | 9 (20.0%) | 616 (68.8%) | **<0.001** |
| Other | 20 (2.1%) | 1 (2.2%) | 19 (2.1%) | >0.900 |

<sup>1</sup>n (%)
<sup>2</sup>Fisher's exact test; Pearson's Chi-squared test

Despite the small proportion of participants declining a new TB vaccine, we performed the 5C composite analysis comparing the groups of those who would accept vaccination with the refusals and unsure individuals. Those who would not take the vaccine had more distrust in vaccine safety (77.8% vs 22.7%, p<0.001), efficacy (55.6% vs 7.8%, p <0.001) and healthcare (60.0% vs 7.9%, p <0.001) than those who would accept vaccination. These individuals declining vaccination also more frequently reported not having adequate sources of information, not considering healthcare workers or government agencies as reliable (24.4% vs 5.5%, p < 0.001). Although both groups had high proportions of people concerned about TB disease, those who would decline the vaccine were less concerned about TB infection or disease severity, as evaluated by the complacency composite variable (77.8% vs 88.8%, p =0.024). There was no difference in the composite evaluation for community coercion between the groups, assessing the answers to family or other PDLs influencing vaccine decision, however, it was reported by a significant proportion of the overall population (13.0%) **(Table 3)**

**Table 3.** Variables associated with not accepting a new proposed TB vaccine among people deprived of liberty in Brazil (n = 941)

| Variables | Overall<br>N = 941<sup>1</sup> | Would you take the TB vaccine? |  | p-value<sup>2</sup> |
| --- | --- | --- | --- | --- |
|  |  | No / Unsure<br>N = 45<sup>1</sup> | Yes<br>N = 896<sup>2</sup> |  |
| ***Confidence*** |  |  |  |  |
| Composite distrust in vaccine safety | 238 (25.3%) | 35 (77.8%) | 203 (22.7%) | **<0.001** |
| Composite distrust in vaccine efficacy | 95 (10.1%) | 25 (55.6%) | 70 (7.8%) | **<0.001** |
| Composite distrust in healthcare system/workers | 98 (10.4%) | 27 (60.0%) | 71 (7.9%) | **<0.001** |
| ***Complacency*** |  |  |  |  |
| Concerned about TB | 831 (88.3%) | 35 (77.8%) | 796 (88.8%) | **0.024** |
| ***Calculation*** |  |  |  |  |
| Bad previous TB knowledge | 388 (41.2%) | 18 (40.0%) | 370 (41.3%) | 0.900 |
| Not adequate sources of information | 60 (6.4%) | 11 (24.4%) | 49 (5.5%) | **<0.001** |
| ***Collective constrains*** |  |  |  |  |
| Reported community coercion | 122 (13.0%) | 9 (20.0%) | 113 (13.0%) | 0.150 |

<sup>1</sup>n (%)
<sup>2</sup>Pearson's Chi-squared test; Fisher's exact test

There were also gender related differences across the responses. In male prison units the reported previous knowledge about TB was lower compared to participants from female prisons (15.3% vs 34.6%, p<0.001). Among the reasons to deny vaccination, male PDL reported more health-related (18.1% vs 3.0%, p<0.001) and community related barriers (8.1% vs 3.0%, p=0.008) than females. Compared to females, male PDL trusted the facility-based healthcare more (82.7% vs 74.8%, p = 0.007) but there was no significant difference in confidence regarding other sources of information **(Supplementary Table 2).** Responses to the Likert scale questions evidenced that females disagree more about statements regarding their concerns of TB disease (19% vs 25%) and severity (23% vs 17%) than males, however overall agreement with vaccine importance was high and similar between males and females (**Supplementary Figure 1**). The 5C composite analysis showed that male PDL have more distrust in vaccine safety (28.9% vs 14.5%, p<0.001), more community coercion regarding vaccine decisions (15.4% vs 5.6%, p<0.001) and reported more insufficient previous TB knowledge (44.6% vs 31.3% p<0.001) compared to females **(Supplementary Table 3),** despite male prisons presenting with a higher prevalence of previous TB and proportion of people reporting contact with TB cases in the same cell.

## Discussion

In this cross-sectional evaluation of six male and two female prisons in Brazil we detected a very high reported acceptance rate for a proposed new TB vaccine. Participants reporting that they would decline a potential new TB vaccine had a higher proportion of reported active smoking and illicit drug use; the main reasons reported for declining it were related to confidence in the vaccine efficacy and safety, as well as distrust of healthcare professionals and government official sources. We also encountered a lower rate of concern about TB disease among those that declined uptake of a new TB vaccine Surprisingly, 22.7% and 7.8% of individuals that accepted vaccination also reported some distrust in vaccine safety and efficacy, respectively. To our knowledge, this is the first study to evaluate acceptance of a possible new TB vaccine in carceral settings.

The high vaccine acceptability in our population of PDL across genders, is compatible with national data for other vaccines. The Brazilian national immunization program has a history of over half a century successfully implementing vaccination campaigns, promoting vaccine propaganda and presenting successful coverage results leading to a scenario of low hesitancy rates in the general population (26,27). Despite reductions in the overall coverage in the last decade due to the surge of anti-vaccine movements and reduction of public investment in healthcare (28), vaccine hesitancy remained low regardless of the increased distrust movement in the initial distributions of COVID-19 vaccines (29).

As evidenced by our findings, higher vaccine hesitation has been reported previously to be associated with illicit drug use and tobacco smoking (30,31). This finding is concerning since our sample evidences a high proportion of reported drug use and that one quarter of the individuals are current active tobacco smokers, this pattern of substance use is even higher in carceral settings worldwide (32). These specific groups can be evaluated in targeted approaches if future vaccine uptake proves to be lower than expected.

We observed elevated rates of confidence issues, even among those who would accept vaccination. Overall, a quarter of PDL have some distrust in vaccine safety and a tenth reported distrust in vaccine efficacy or healthcare system and professionals. Distrust in these factors is a strong determinant of vaccine acceptability (19,33); previous surveys conducted in the overall Brazilian population reported that confidence issues were the most relevant motives among people who hesitated in vaccination, with 41.4% and 25.5% of those did not believing that vaccines were safe or effective, respectively (15).

Moreover, in PDL there has historically been ethic abuses, neglect and insufficient access to healthcare take place (20,34). Recently there has been frequent discussions regarding the inclusion of PDL in vaccine trials (34). Previous qualitative research performed in some of the prisons included in this study support these findings and present solutions proposed by the PDL interviewed; increasing vaccine education could strengthen consent and ensuring PDL participation in study oversight with community counsels could promote trust within the population (20). We hypothesize that this community engagement can increase vaccine acceptability when vaccination campaigns start.

Vaccine education efforts should also be aligned with TB disease education. Previous studies with COVID-19 have also suggested that education about disease could improve vaccine uptake in prisons (35,36). Our findings highlighted that more concerns about TB and previously knowing someone with TB were positively associated with vaccine acceptance. Willingness to participate in TB vaccine trials has been associated with disease knowledge in other contexts (37), finding that could also represent a possible association with vaccine acceptance since PDL report fear of TB disease for personal or second hand experience as factors influencing vaccination choices (20). In prison settings, the high incidence of TB (7) leads to the high proportion of individuals reporting cellmates with TB and knowing someone with a TB diagnosis in the past; however, this contact does not guarantee a good understanding of the disease and personal risks.

Although only a small proportion of all participants reported feeling community coercion influencing their decision, this perception was substantially more common among those who declined vaccination. Future trials and vaccine campaigns must ensure that there are measures in place to respect individual autonomy of decision (34).

Our study has several limitations. Our primary outcome, TB vaccine acceptability, is concerning a hypothetical product that it is not currently available. Within the vaccine approval pipeline unexpected factors such as considerable side effects, efficacy of the vaccine, inadequate safety information or distrust in the approval process might reduce the expected acceptability significantly. Also, there is no standardized measurements or questionnaires for hypothetical vaccination scenarios (38), most hesitancy studies are regarding existing products or compare groups that already were vaccinated with those who refused.

Finally, despite evaluating several prison units in all regions of Brazil, these results might not reflect acceptability in the overall population or PDL in other countries. The burden of TB in prisons, especially in Brazil, is disproportional which might lead to a higher vaccine acceptance due to increased knowledge and concern about the disease. Also, due to social desirability bias, it is possible that participants may have falsely reported that they would accept a new TB vaccine due to fear of conflict or exclusion within the prison community for not accepting interventions that would be beneficial for the broader prison population.

## Conclusion

In conclusion, we identified that most incarcerated individuals in our sample would accept a proposed new TB vaccine. Younger age, tobacco smoking and illicit drug use were associated with lower acceptability. Individuals that refused vaccination reported more often distrust in vaccine safety, efficacy and healthcare system, were less concerned about TB disease and had more inadequate vaccine information sources. Our findings indicate that a new vaccine would be well accepted overall among PDL and that prospective vaccination projects should aim to increase populational trust if adequate acceptance is not reached.

## Data Availability

The dataset used for the present study are available upon reasonable request to the corresponding author.

## Declarations

### Consent for publication

Study participants were consented to have their de-identified data used for scientific purposes including the publication of this manuscript.

## Authors’ contributions

RGW and JC conceived the study design. JVBB, GGAA, KMS and MSV participated in the data collection process. JVBB, GGAA, KMS and MSV assisted with the sample collections/processing and data collection process. JVBB, KAT and RAC performed the analyses. JVBB, RAC and KAT interpreted the results. JVBB and GGAA wrote the first draft of the manuscript. RAC and KAT critically revised the first draft of the manuscript. KMS and MSV assisted with further drafting and revisions of the manuscript. All authors reviewed and approved the final version of the manuscript.

## SUPPLEMENTARY TABLES & FIGURES

**Supplementary Table 1.** Characteristics of incarcerated individuals included in the vaccine acceptability study, by willingness to take a proposed TB vaccine (n = 941)

| Variables | Overall N = 941<sup>1</sup> | Would you take the vaccine? |  | p-value<sup>2</sup> |
| --- | --- | --- | --- | --- |
|  |  | No / Unsure N = 45<sup>1</sup> | Yes N = 896<sup>1</sup> |  |
| Sociodemographic characteristics |  |  |  |  |
| Gender |  |  |  | 0.004 |
| Male | 707 (75.1%) | 42 (93.3%) | 665 (74.2%) |  |
| Female | 234 (24.9%) | 3 (6.7%) | 231 (25.8%) |  |
| Age categories |  |  |  | 0.073 |
| 18 – 25 years old | 166 (17.6%) | 15 (33.3%) | 151 (16.9%) |  |
| 26 – 35 years old | 421 (44.7%) | 16 (35.6%) | 405 (44.7%) |  |
| 36 – 45 years old | 250 (26.6%) | 10 (22.2%) | 240 (26.6%) |  |
| Above 45 years old | 104 (11.1%) | 4 (8.9%) | 100 (11.1%) |  |
| Race |  |  |  | 0.500 |
| White | 185 (19.7%) | 11 (24.4%) | 174 (19.4%) |  |
| Black | 192 (20.4%) | 8 (17.8%) | 184 (20.5%) |  |
| Mixed | 546 (58.0%) | 25 (55.6%) | 521 (58.1%) |  |
| Yellow | 10 (1.1%) | 0 (0%) | 10 (1.1%) |  |
| Indigenous | 8 (0.9%) | 1 (2.2%) | 7 (0.8%) |  |
| Marital Status |  |  |  | 0.100 |
| Divorced | 42 (4.5%) | 0 (0%) | 42 (4.7%) |  |
| Married | 209 (22.2%) | 17 (37.8%) | 192 (21.4%) |  |
| Single | 559 (59.4%) | 22 (48.9%) | 537 (59.9%) |  |
| Stable Union | 123 (13.1%) | 6 (13.3%) | 117 (13.1%) |  |
| Widowed | 8 (0.9%) | 0 (0%) | 8 (0.9%) |  |
| Schooling |  |  |  | 0.700 |
| Incomplete basic education | 455 (48.4%) | 23 (51.1%) | 432 (48.2%) |  |
| Complete basic education or higher | 486 (51.6%) | 22 (48.9%) | 464 (51.8%) |  |
| Family income |  |  |  | >0.900 |
| Less then 1 minimum wage | 164 (17.4%) | 9 (20.0%) | 155 (17.3%) |  |
| 1-2 minimum wages | 422 (44.8%) | 18 (40.0%) | 404 (45.1%) |  |
| 3-4 minimum wages | 74 (7.9%) | 3 (6.7%) | 71 (7.9%) |  |
| >4 minimum wages | 47 (5.0%) | 3 (6.7%) | 44 (4.9%) |  |
| Not informed | 234 (24.9%) | 12 (26.7%) | 222 (24.8%) |  |
| Prison characteristics |  |  |  |  |
| Previously incarcerated | 677 (71.9%) | 32 (71.1%) | 645 (72.0%) | 0.900 |
| Previous contact with TB case in the same cell | 323 (34.3%) | 19 (42.2%) | 304 (33.9%) | 0.300 |
| Current incarceration time |  |  |  | 0.400 |
| <1 year | 357 (37.9%) | 14 (31.1%) | 343 (38.3%) |  |
| 1-2 years | 275 (29.2%) | 18 (40.0%) | 257 (28.7%) |  |
| 3-4 years | 127 (13.5%) | 5 (11.1%) | 122 (13.6%) |  |
| 5 years or above | 182 (19.3%) | 8 (17.8%) | 174 (19.4%) |  |
| Clinical characteristics |  |  |  |  |
| Previous TB history | 87 (9.2%) | 5 (11.1%) | 82 (9.2%) | 0.600 |
| Living with HIV | 13 (1.4%) | 0 (0%) | 13 (1.5%) | >0.900 |
| Alcohol abuse | 42 (4.5%) | 4 (8.9%) | 38 (4.2%) | 0.140 |
| Current tobacco smoking | 243 (25.8%) | 20 (44.4%) | 223 (24.9%) | 0.003 |
| Illicit drug use | 144 (15.3%) | 12 (26.7%) | 132 (14.7%) | 0.030 |

<sup>1</sup>n (%); Median (Q1 - Q3)
<sup>2</sup>Pearson's Chi-squared test; Wilcoxon rank sum test; Fisher's exact test

**Supplementary Table 2.**
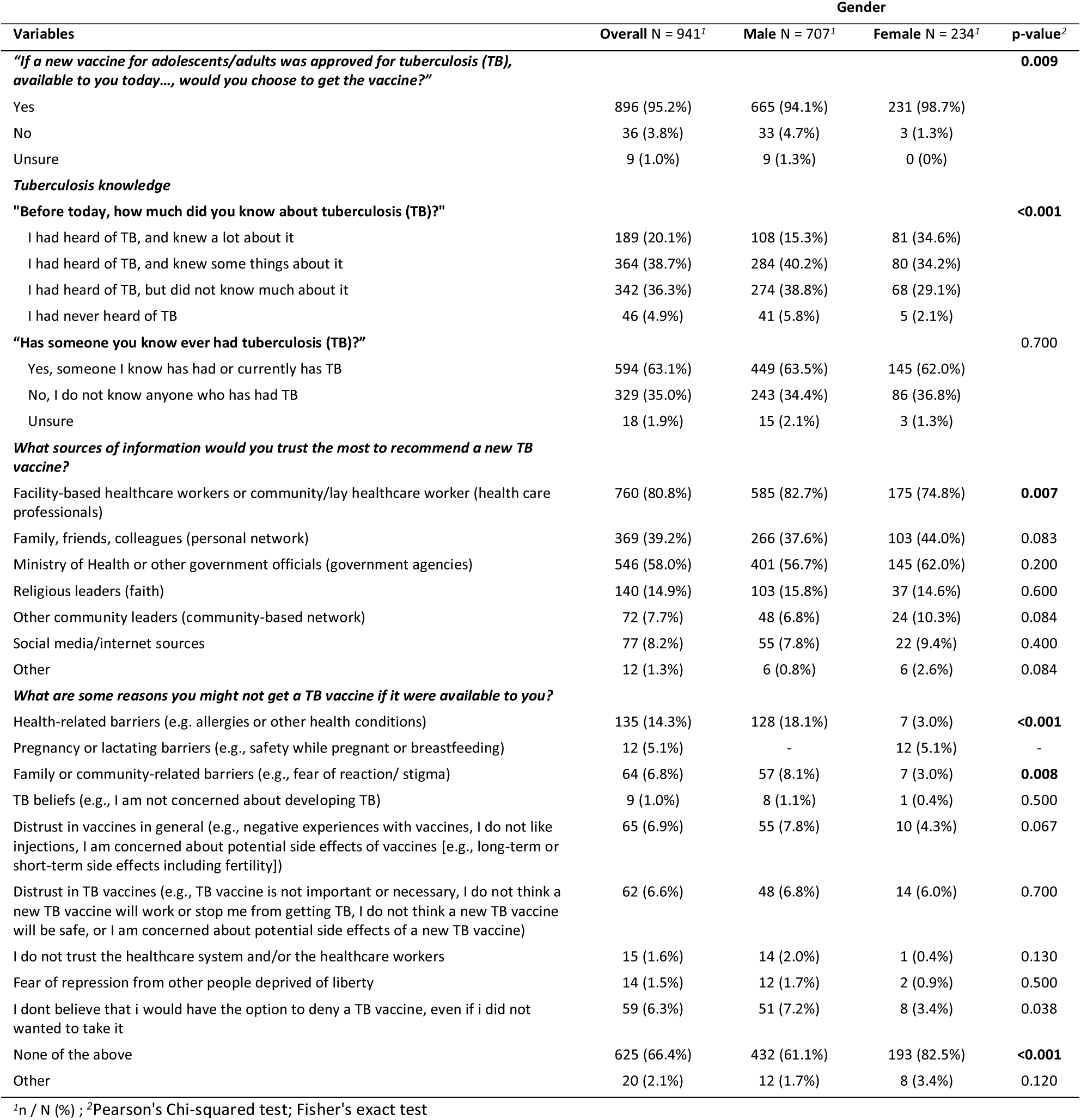
Vaccine acceptability questions assessing knowledge, information sources and reasons to not get vaccinated in people deprived of liberty, by gender (n = 941)

| Variables | Gender |  |  | p-value<sup>2</sup> |
| --- | --- | --- | --- | --- |
|  | Overall N = 941<sup>1</sup> | Male N = 707<sup>1</sup> | Female N = 234<sup>1</sup> |  |
| ***"If a new vaccine for adolescents/adults was approved for tuberculosis (TB), available to you today..., would you choose to get the vaccine?"*** |  |  |  | **0.009** |
| Yes | 896 (95.2%) | 665 (94.1%) | 231 (98.7%) |  |
| No | 36 (3.8%) | 33 (4.7%) | 3 (1.3%) |  |
| Unsure | 9 (1.0%) | 9 (1.3%) | 0 (0%) |  |
| ***Tuberculosis knowledge*** |  |  |  |  |
| ***"Before today, how much did you know about tuberculosis (TB)?"*** |  |  |  | **<0.001** |
| I had heard of TB, and knew a lot about it | 189 (20.1%) | 108 (15.3%) | 81 (34.6%) |  |
| I had heard of TB, and knew some things about it | 364 (38.7%) | 284 (40.2%) | 80 (34.2%) |  |
| I had heard of TB, but did not know much about it | 342 (36.3%) | 274 (38.8%) | 68 (29.1%) |  |
| I had never heard of TB | 46 (4.9%) | 41 (5.8%) | 5 (2.1%) |  |
| ***"Has someone you know ever had tuberculosis (TB)?"*** |  |  |  | 0.700 |
| Yes, someone I know has had or currently has TB | 594 (63.1%) | 449 (63.5%) | 145 (62.0%) |  |
| No, I do not know anyone who has had TB | 329 (35.0%) | 243 (34.4%) | 86 (36.8%) |  |
| Unsure | 18 (1.9%) | 15 (2.1%) | 3 (1.3%) |  |
| ***What sources of information would you trust the most to recommend a new TB vaccine?*** |  |  |  |  |
| Facility-based healthcare workers or community/lay healthcare worker (health care professionals) | 760 (80.8%) | 585 (82.7%) | 175 (74.8%) | **0.007** |
| Family, friends, colleagues (personal network) | 369 (39.2%) | 266 (37.6%) | 103 (44.0%) | 0.083 |
| Ministry of Health or other government officials (government agencies) | 546 (58.0%) | 401 (56.7%) | 145 (62.0%) | 0.200 |
| Religious leaders (faith) | 140 (14.9%) | 103 (15.8%) | 37 (14.6%) | 0.600 |
| Other community leaders (community-based network) | 72 (7.7%) | 48 (6.8%) | 24 (10.3%) | 0.084 |
| Social media/internet sources | 77 (8.2%) | 55 (7.8%) | 22 (9.4%) | 0.400 |
| Other | 12 (1.3%) | 6 (0.8%) | 6 (2.6%) | 0.084 |
| ***What are some reasons you might not get a TB vaccine if it were available to you?*** |  |  |  |  |
| Health-related barriers (e.g. allergies or other health conditions) | 135 (14.3%) | 128 (18.1%) | 7 (3.0%) | **<0.001** |
| Pregnancy or lactating barriers (e.g., safety while pregnant or breastfeeding) | 12 (5.1%) | - | 12 (5.1%) | - |
| Family or community-related barriers (e.g., fear of reaction/ stigma) | 64 (6.8%) | 57 (8.1%) | 7 (3.0%) | **0.008** |
| TB beliefs (e.g., I am not concerned about developing TB) | 9 (1.0%) | 8 (1.1%) | 1 (0.4%) | 0.500 |
| Distrust in vaccines in general (e.g., negative experiences with vaccines, I do not like injections, I am concerned about potential side effects of vaccines [e.g., long-term or short-term side effects including fertility]) | 65 (6.9%) | 55 (7.8%) | 10 (4.3%) | 0.067 |
| Distrust in TB vaccines (e.g., TB vaccine is not important or necessary, I do not think a new TB vaccine will work or stop me from getting TB, I do not think a new TB vaccine will be safe, or I am concerned about potential side effects of a new TB vaccine) | 62 (6.6%) | 48 (6.8%) | 14 (6.0%) | 0.700 |
| I do not trust the healthcare system and/or the healthcare workers | 15 (1.6%) | 14 (2.0%) | 1 (0.4%) | 0.130 |
| Fear of repression from other people deprived of liberty | 14 (1.5%) | 12 (1.7%) | 2 (0.9%) | 0.500 |
| I don't believe that I would have the option to deny a TB vaccine, even if I did not want to take it | 59 (6.3%) | 51 (7.2%) | 8 (3.4%) | 0.038 |
| None of the above | 625 (66.4%) | 432 (61.1%) | 193 (82.5%) | **<0.001** |
| Other | 20 (2.1%) | 12 (1.7%) | 8 (3.4%) | 0.120 |

<sup>1</sup>n / N (%); <sup>2</sup>Pearson's Chi-squared test; Fisher's exact test

**Supplementary Figure 1.**
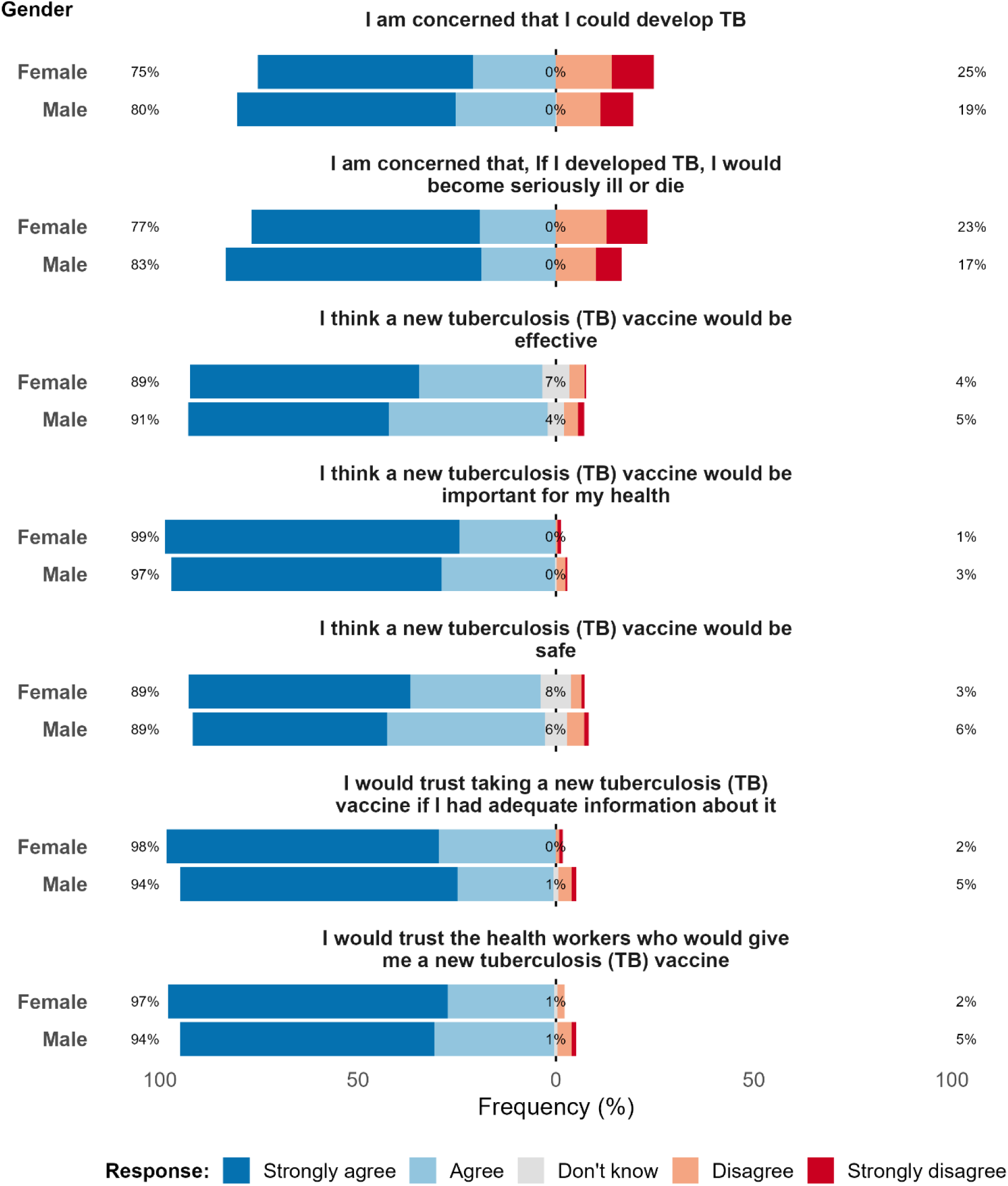
Likert scale responses to questions related to tuberculosis concerns and vaccine acceptability among people deprived of liberty, stratified by gender (N = 941)

**Supplementary Table 3.** Variables associated with not accepting the new proposed TB vaccine among people deprived of liberty in Brazil, by gender (n = 941)

| Variables | Overall N = 941<sup>1</sup> | Gender |  | p-value<sup>2</sup> |
| --- | --- | --- | --- | --- |
|  |  | Male N = 707<sup>1</sup> | Female N = 234<sup>1</sup> |  |
| **Confidence** |  |  |  |  |
| Composite distrust in vaccine safety | 238 (25.3%) | 204 (28.9%) | 34 (14.5%) | <0.001 |
| Composite distrust in vaccine efficacy | 95 (10.1%) | 73 (10.3%) | 22 (9.4%) | 0.700 |
| Composite distrust in healthcare system/workers | 98 (10.4%) | 77 (10.9%) | 21 (9.0%) | 0.400 |
| **Complacency** |  |  |  |  |
| Concerned about TB | 831 (88.3%) | 628 (88.8%) | 203 (86.8%) | 0.400 |
| **Calculation** |  |  |  |  |
| Bad previous TB knowledge | 388 (41.2%) | 315 (44.6%) | 73 (31.2%) | <0.001 |
| Not adequate sources of information | 60 (6.4%) | 44 (6.2%) | 16 (6.8%) | 0.700 |
| **Collective constrains** |  |  |  |  |
| Reported community coercion | 122 (13.0%) | 109 (15.4%) | 13 (5.6%) | <0.001 |

<sup>1</sup>n (%)
<sup>2</sup>Pearson's Chi-squared test; Fisher's exact test

## References

1. Global tuberculosis report 2025 [Internet]. 1st ed. Geneva: World Health Organization; 2025 [cited 2025 Nov 30]. Available from: https://www.who.int/publications/i/item/9789240116924

2. Implementing the end TB strategy: the essentials, 2022 update. 1st ed. Geneva: World Health Organization; 2022.

3. An Investment Case for New Tuberculosis Vaccines. 1st ed. Geneva: World Health Organization; 2022. 1 p.

4. TB Vaccine Clinical Pipeline [Internet]. Working Group on New TB Vaccines. [cited 2025 Oct 9]. Available from: https://newtbvaccines.org/tb-vaccine-pipeline/clinical-phase/

5. Luabeya AKK, Rozot V, Imbratta C, Ratangee F, Shenje J, Tameris M, et al. Live-attenuated *Mycobacterium tuberculosis* vaccine, MTBVAC, in adults with or without *M tuberculosis* sensitisation: a single-centre, phase 1b–2a, double-blind, dose-escalation, randomised controlled trial. The Lancet Global Health. 2025 Jun 1;13(6):e1030–42.

6. Dagnew AF, Han LL, Naidoo K, Fairlie L, Innes JC, Middelkoop K, et al. Safety and immunogenicity of investigational tuberculosis vaccine M72/AS01E–4 in people living with HIV in South Africa: an observer-blinded, randomised, controlled, phase 2 trial. The Lancet HIV. 2025 Aug 1;12(8):e546–55.

7. Cords O, Martinez L, Warren JL, O’Marr JM, Walter KS, Cohen T, et al. Incidence and prevalence of tuberculosis in incarcerated populations: a systematic review and meta-analysis. Lancet Public Health. 2021 May;6(5):e300–8.

8. Mabud TS, de Lourdes Delgado Alves M, Ko AI, Basu S, Walter KS, Cohen T, et al. Evaluating strategies for control of tuberculosis in prisons and prevention of spillover into communities: An observational and modeling study from Brazil. PLoS Med. 2019 Jan;16(1):e1002737.

9. Sacchi FPC, Praça RM, Tatara MB, Simonsen V, Ferrazoli L, Croda MG, et al. Prisons as Reservoir for Community Transmission of Tuberculosis, Brazil. Emerg Infect Dis. 2015 Mar;21(3):452–5.

10. Liu YE, Mabene Y, Camelo S, Rueda ZV, Pelissari DM, Dockhorn Costa Johansen F, et al. Mass incarceration as a driver of the tuberculosis epidemic in Latin America and projected effects of policy alternatives: a mathematical modelling study. Lancet Public Health. 2024 Nov;9(11):e841–51.

11. Ranzani OT, Pescarini JM, Martinez L, Garcia-Basteiro AL. Increasing tuberculosis burden in Latin America: an alarming trend for global control efforts. BMJ Glob Health [Internet]. 2021 Mar 24 [cited 2025 Oct 10];6(3). Available from: https://gh.bmj.com/content/6/3/e005639

12. Madeddu G, Vroling H, Oordt-Speets A, Babudieri S, O’Moore É, Noordegraaf MV, et al. Vaccinations in prison settings: A systematic review to assess the situation in EU/EEA countries and in other high income countries. Vaccine. 2019 Aug 14;37(35):4906–19.

13. Moazen B, Ismail N, Agbaria N, Mazzilli S, Petri D, Amaya A, et al. Vaccination against emerging and reemerging infectious diseases in places of detention: a global multistage scoping review. Front Public Health. 2024 Jan 29;12:1323195.

14. Ismail N, Tavoschi L, Moazen B, Roselló A, Plugge E. COVID-19 vaccine for people who live and work in prisons worldwide: A scoping review. PLOS ONE. 2022 de set. de;17(9):e0267070.

15. Brown AL, Sperandio M, Turssi CP, Leite RMA, Berton VF, Succi RM, et al. Vaccine confidence and hesitancy in Brazil. Cad Saúde Pública. 2018;34:e00011618.

16. Sato APS. What is the importance of vaccine hesitancy in the drop of vaccination coverage in Brazil? Revista de Saúde Pública. 2018 Nov 22;52:96–96.

17. Agência Brasil [Internet]. 2022 [cited 2025 Oct 10]. Unidades socioeducativas chegam a 97% de vacinação contra a covid. Available from: https://agenciabrasil.ebc.com.br/radioagencia-nacional/saude/audio/2022-10/unidades-socioeducativas-chegam-97-de-vacinacao-contra-covid

18. Rocha AC da S, Santos AAP dos, Lucena TS de, Santos WB dos, Tavares NV da S, Moraes MM de. Análise da vacinação contra Covid-19 na população privada de liberdade: estudo ecológico. Revista Enfermagem UERJ. 2024 May 29;32:e76740–e76740.

19. MacDonald NE. Vaccine hesitancy: Definition, scope and determinants. Vaccine. 2015 Aug 14;33(34):4161–4.

20. Pires MCCF, Liu YE, Lemos EF, Silva LF da, Croda MG, Magalhães M, et al. Perceptions of persons deprived of liberty regarding tuberculosis vaccine research [Internet]. medRxiv; 2025 [cited 2025 Oct 10]. p. 2025.07.04.25330885. Available from: https://www.medrxiv.org/content/10.1101/2025.07.04.25330885v1

21. Larouzé B, Ventura M, Sánchez AR, Diuana V. Tuberculose nos presídios brasileiros: entre a responsabilização estatal e a dupla penalização dos detentos. Cad Saúde Pública. 2015;31:1127–30.

22. Vandergrift LA, Christopher PP. Do prisoners trust the healthcare system? Health & Justice. 2021 Jul 3;9(1):15.

23. Betsch C, Schmid P, Heinemeier D, Korn L, Holtmann C, Böhm R. Beyond confidence: Development of a measure assessing the 5C psychological antecedents of vaccination. PLoS One. 2018 Dec 7;13(12):e0208601.

24. R Core Team. R: A language and environment for statistical computing. R Foundation for Statistical Computing;

25. Boletim Epidemiológico - HIV e Aids (2024) — Departamento de HIV, Aids, Tuberculose, Hepatites Virais e Infecções Sexualmente Transmissíveis [Internet]. [cited 2025 Feb 23]. Available from: https://www.gov.br/aids/pt-br/central-de-conteudo/boletins-epidemiologicos/2024/boletim_hiv_aids_2024e.pdf/view

26. Minakawa MM, Frazão P. The Trajectory of Brazilian Immunization Program between 1980 and 2018: From the Virtuous Cycle to the Vaccine Coverage Decline. Vaccines (Basel). 2023 Jul 1;11(7):1189.

27. Pércio J, Fernandes EG, Maciel EL, de Lima NVT. 50 years of the Brazilian National Immunization Program and the Immunization Agenda 2030. Epidemiol Serv Saude. 32(3):e20231009.

28. Césare N, Mota TF, Lopes FFL, Lima ACM, Luzardo R, Quintanilha LF, et al. Longitudinal profiling of the vaccination coverage in Brazil reveals a recent change in the patterns hallmarked by differential reduction across regions. Int J Infect Dis. 2020 Sep;98:275–80.

29. Moore DCBC, Nehab MF, Camacho KG, Reis AT, Junqueira-Marinho M de F, Abramov DM, et al. Low COVID-19 vaccine hesitancy in Brazil. Vaccine. 2021 Oct 8;39(42):6262–8.

30. Havelka EM, Sanfilippo JE, Juneau PL, Sherman G, Cooper D, Leggio L. The effect of alcohol, tobacco, and other drug use on vaccine acceptance, uptake, and adherence: a systematic review. Alcohol Alcohol. 2024 Oct 5;59(6):agae057.

31. Jackson SE, Paul E, Brown J, Steptoe A, Fancourt D. Negative Vaccine Attitudes and Intentions to Vaccinate Against Covid-19 in Relation to Smoking Status: A Population Survey of UK Adults. Nicotine Tob Res. 2021 Mar 5;23(9):1623–8.

32. Favril L, Rich JD, Hard J, Fazel S. Mental and physical health morbidity among people in prisons: an umbrella review. Lancet Public Health. 2024 Mar 27;9(4):e250–60.

33. Pereira ET, Iasulaitis S, Greco BC. Analysis of causal relations between vaccine hesitancy for COVID-19 vaccines and ideological orientations in Brazil. Vaccine. 2024 May 10;42(13):3263–71.

34. Andrews JR, Charalambous S, Churchyard G, Cobelens F, Fernández-Escobar C, Frick M, et al. The participation of people deprived of liberty in tuberculosis vaccine trials: should they be protected from research, or through research? The Lancet Infectious Diseases [Internet]. 2025 Jun 27 [cited 2025 Oct 2]; Available from: https://www.sciencedirect.com/science/article/pii/S1473309925003056

35. Ortiz-Paredes D, Varsaneux O, Worthington J, Park H, MacDonald SE, Basta NE, et al. Reasons for COVID-19 vaccine refusal among people incarcerated in Canadian federal prisons. PLoS One. 2022 Mar 9;17(3):e0264145.

36. Lessard D, Ortiz-Paredes D, Park H, Varsaneux O, Worthington J, Basta NE, et al. Barriers and facilitators to COVID-19 vaccine acceptability among people incarcerated in Canadian federal prisons: A qualitative study. Vaccine X. 2022 Feb 19;10:100150.

37. Shu E, Sobieszczyk ME, Sal y Rosas VG, Segura P, Galea JT, Lecca L, et al. Knowledge of tuberculosis and vaccine trial preparedness in Lima, Peru. The International Journal of Tuberculosis and Lung Disease. 2017 Dec 1;21(12):1288–93.

38. Larson HJ, Jarrett C, Schulz WS, Chaudhuri M, Zhou Y, Dube E, et al. Measuring vaccine hesitancy: The development of a survey tool. Vaccine. 2015 Aug 14;33(34):4165–75.

