## Supplementary materials, Appendix 1 for "Acceptability of a proposed new tuberculosis vaccine among people deprived of liberty in Brazil"

**SUPPLEMENTARY MATERIAL – APPENDIX 1**

**VACCINE ACCEPTABILITY QUESTIONNAIRE**

1. Translated vaccine acceptability questionnaire in English---------------------------------------------- Page 2

**English translation of the original vaccine questionnaire**

**A. TB knowledge overall**

1. Before today, how much did you know about tuberculosis (TB)?

⚪ I had heard of TB, and knew a lot about it

⚪ I had heard of TB, and knew some things about it

⚪ I had heard of TB, but did not know much about it

⚪ I had never heard of TB

2. To your knowledge, have you ever been treated for tuberculosis (TB) disease in the past?

⚪ Yes, I have previously been treated for TB disease

⚪ No, I have never been treated for TB disease

⚪ I would rather not discuss it

3. Has someone you know ever had tuberculosis (TB)?

⚪ Yes, someone I know has had or currently has TB

⚪ No, I do not know anyone who has had TB

⚪ Unsure

4. How strongly do you agree or disagree with the following affirmations?

|  | Strongly agree | Agree | Disagree | Strongly disagree | Don’t know |
| --- | --- | --- | --- | --- | --- |
| I am concerned that I could develop TB |  |  |  |  |  |
| If I developed TB, I am concerned that I would become seriously ill or die |  |  |  |  |  |

**B. TB vaccines**

5. If a new vaccine for adolescents/adults was approved for tuberculosis (TB) and recommended by the Brazil Ministry of Health, how strongly do you agree or disagree with the following affirmations?

|  | Strongly agree | Agree | Disagree | Strongly disagree | Don’t know |
| --- | --- | --- | --- | --- | --- |
| I think a new tuberculosis (TB) vaccine would be important for my health |  |  |  |  |  |
| I think a new tuberculosis (TB) vaccine would be safe |  |  |  |  |  |
| I think a new tuberculosis (TB) vaccine would be effective |  |  |  |  |  |
| I would trust the health workers who would give me a new tuberculosis (TB) vaccine |  |  |  |  |  |
| I would trust taking a new tuberculosis (TB) vaccine if I had adequate information about it |  |  |  |  |  |

6. If a new vaccine for adolescents/adults was approved for tuberculosis (TB), available to you today, recommended by the Brazil Ministry of Health, and your healthcare provider recommended it, would you choose to get the vaccine?

⚪ Yes (go to question 8)

⚪ No (go to question 8)

⚪ Unsure (awnser to question 7)

7. *[If “Unsure” in question 6]* If a new tuberculosis (TB) vaccine was available today, and you had to make a choice, would you choose to get the vaccine?

⚪ Yes

⚪ No

8. What sources of information would you trust the most to recommend a new TB vaccine? Please, select all applicable options. Please, do not answer until all responses have been read.

⬜ Facility-based healthcare workers or community/lay healthcare worker (health care professionals)

⬜ Family, friends, colleagues (personal network)

⬜ Ministry of Health or other government officials (government agencies)

⬜ Religious leaders (faith)

⬜ Other community leaders (community-based network)

⬜ Social media/internet sources

⬜Other _________________________________________________________________________________

9. What are some reasons you might not get a TB vaccine if it were available to you? Please, select all applicable options. Please, do not answer until all responses have been read.

⬜ Health-related barriers (e.g., I have a prior history of allergies or other health conditions that may make certain vaccines unsafe)

⬜ Pregnancy or lactating barriers (e.g., I have concerns about the safety of receiving a vaccine while pregnant or breastfeeding)

⬜ Family or community-related barriers (e.g., I would be concerned about the reaction or objections from my spouse, family and/or the community, my religion prohibits or discourages receiving vaccines, stigma)

⬜ TB beliefs (e.g., I am not concerned about developing TB)

⬜ Distrust in vaccines in general (e.g., I have had previous negative experiences with vaccines, I do not like injections, I am concerned about potential side effects of vaccines [e.g., long-term or short-term side effects including fertility])

⬜ Distrust in TB vaccines (e.g., I do not think a new TB vaccine is important or necessary, I do not think a new TB vaccine will work or stop me from getting TB, I do not think a new TB vaccine will be safe, or I am concerned about potential side effects of a new TB vaccine [e.g., long-term or short-term side effects including fertility])

⬜ I do not trust the healthcare system and/or the healthcare workers

⬜ Fear of repression from other people deprived of liberty

⬜ I don’t believe that I would have the option to deny a TB vaccine, even if i did not wanted to take it

⬜ None of the above

⬜Other ____________________________________________________________________________________
